# HLA and red blood cell antigen genotyping in SARS-CoV-2 convalescent plasma donors

**DOI:** 10.1101/2021.03.17.21253821

**Authors:** William Lemieux, Josée Perreault, Gabriel André Leiva-Torres, Nadia Baillargeon, Jessica Constanzo Yanez, Marie-Claire Chevrier, Lucie Richard, Antoine Lewin, Patrick Trépanier

## Abstract

**Introduction:** The SARS-CoV-2 pandemic has put significant additional pressure on healthcare systems throughout the world. The identification of at-risk population beyond age, pre-existing medical conditions and socioeconomic status has been the subject of only a small part of the global COVID-19 research so far. To this day, more data is required regarding the association between HLA allele and red blood cell (RBC) antigens’ expression in regard to SARS-CoV-2 infection susceptibility and virus clearance capability, and COVID-19 susceptibility, severity, and duration.

**Methods:** The phenotypes for ABO and RhD, and the genotypes for 37 RBC antigens and HLA-A, B, C, DRB1, DQB1 and DPB1 were determined using high throughput platforms (Luminex and Next-generation Sequencing) in 90 Caucasian convalescent plasma donors. The results were compared to expected reference frequencies, local and international databases, and literature.

**Results:** The AB group was significantly increased (1.5x, p=0.018) and a non-significant (2.2x, p=0.030) increase was observed for the FY*A allele frequency in the convalescent cohort (N=90) compared to reference frequencies. Some HLA alleles were found significantly overrepresented (HLA-B*44:02, C*05:01, DPB1*04:01, DRB1*04:01 and DRB1*07:01) or underrepresented (A*01:01, B51:01 and DPB1*04:02) in convalescent individuals compared to the local bone marrow registry population.

**Conclusion:** Our study of infection-susceptible but non-hospitalized Caucasian COVID-19 patients contributes to the global understanding of host genetic factors associated with SARS-CoV-2 infection susceptibility and severity of the associated disease.

## Introduction

The coronavirus disease 2019 (COVID-19) caused by the severe acute respiratory syndrome coronavirus 2 (SARS-CoV-2) pandemic has caused over 5.3 million deaths worldwide as of December 2021 (https://covid19.who.int/). Intense work has been done since the beginning of the pandemic to protect the most at-risk populations. Being male, of older age, obese, of a certain ethnic origin, having diabetes, asthma, or many other medical conditions, have all been associated with an increased risk of COVID-19-related complications or death^1–5^. Genetic factors, such as the expression of angiotensin-converting enzyme (ACE)-related genes, may also play a role in disease severity and could serve as a predictive marker for at-risk populations^6^. Efforts in getting a better understanding of the molecular gateways for viral entry and interactions with the host’s response have been relentless, focusing on ACE2 expression, HLA, cytokine storms and TLRs^7^. Such research could lead to an improved understanding of the disease’s susceptibility and severity, of the capacity for virus clearance, of long term symptoms^8^ and mortality consequent to infection in humans.

Relationships between ABO blood group expression and susceptibility to infection by SARS-CoV-2, as well as the risks of hospitalization and death from COVID-19 have been explored by a few groups and recently reviewed^9,10^. O group individuals were identified as having a decreased risk of infection compared to other ABO groups, although no differences were observed regarding hospitalization and death rates associated with COVID-19^11,12^. Several hypotheses to explain this observed link between ABO type and risk of infection have been suggested, such as the presence of anti-A and -B antibodies in O individuals^13^ and the binding of Receptor-Binding Domain (RBD) of the viral Spike protein to group A antigens^14^. Given the important immunological deregulation and potential cytokine storm associated with mortality in COVID-19 patients^15^ and the interesting report from an Italian laboratory of higher rates of direct antiglobulin test (DAT) reactivity in COVID-19 patients^16^, RBC antigens and related antibodies could be involved in COVID-19 and its clinical manifestations.

Of additional importance is the potential contribution of HLAs, given their central role in regulating immunity against viruses^17^. Considerable efforts have been made in trying to predict and identify protective or susceptibility-enhancing HLA alleles^18,19^. Binding assays have shown that HLA-A*02:01 and B*40:01 could preferentially associate with SARS-CoV-2 epitopes^20^. Using bioinformatic prediction, HLA-A*02:03 and A*31:01 were identified as protective, while A*03:02 was identified as a risk allele^21^. *In silico* binding affinity studies have shown that HLA-B*46:01 could increase susceptibility to disease, whereas HLA-B*15:03 is associated with protective immunity, and HLA-A*02:02, B*15:03, C*12:03 were the most frequently encountered haplotypes associated with presentation of viral epitopes^18^. Other retrospective studies aimed at identifying potentially protective or risk-increasing alleles^22–28^ have been published. Such a global effort requires several geographically diverse laboratories to analyze and share available data in order to generate a comprehensive overview.

In the present study, we hypothesized that associations might exist between certain HLA alleles or blood groups and SARS-CoV-2 susceptibility, as well as the ability to overcome infection without hospitalization, in convalescent plasma donors. We therefore sought to analyze and determine the existence of any trends in the expression of an extended panel of RBC antigens (ABO, RhD and 37 other antigens), and of HLA-A, B, C, DRB1, DQB1, and DPB1 alleles within a Caucasian convalescent plasma donor cohort, in comparison to different reference frequencies (textbook, local and international databases, and literature). The identification of differential patterns of RBC antigen or HLA expression in convalescent individuals, who were infected but were not hospitalized, could contribute to a better understanding of SARS-CoV-2 susceptibility and COVID-19 severity, and may also help interpret the discrepancies observed in the rates of infection in different ethnic groups^5,29^.

## Material and methods

### Samples

Caucasian convalescent plasmas were randomly chosen from adult participants of the Québec cohort in the CONCOR-1 clinical trial (#NCT04348656). All subjects received an official diagnosis of COVID-19 by the Québec Provincial Health Authority after epidemiologic investigation or after confirmation by polymerase chain reaction (PCR) test. All subjects were COVID-19 symptomatic during infection, cleared the infection without hospitalization, and were free of symptoms for at least two weeks before donation. Since COVID-19 diagnostic was given before spring 2021, subjects were presumably infected with the Alpha (B.1.1.7) and Beta (B.1.351) variant, since no case of the Delta (B.1.617.2) variant was yet reported in the province of Quebec^30^. All subjects were self-identified Caucasians. Our convalescent plasma donor cohort had an average age of 40.4 ± 15.0 years and consisted of 68% males. Donors were not selected with respect to their ABO group. All donors gave consent to participate in this research project, which was approved by the Héma-Québec Research Ethics Committee.

### Phenotyping and genotyping

ABO and RhD phenotype testing were done by serologic detection using the PK7300 from Beckman Coulter, as per the manufacturer’s protocol. The DNA used for genotyping was extracted from the buffy coat of whole blood samples collected in ethylenediaminetetraacetic acid (EDTA) tubes, using QIAamp Blood Mini kit (Qiagen, Hilden, Germany). RBC genotyping was done using the Luminex xMAP® technology with the ID CORE XT platform (Progenika Biopharma-Grifols, Bizkaia, Spain), as per the manufacturer’s protocol, for the following blood group antigens: Rh (C, c, E, e, C^w^, hr^S^, hr^B^, V, VS), Kell (K, k, Kp^a,^ Kp^b^, Js^a,^ Js^b^), Kidd (Jk^a^, Jk^b^), Duffy (Fy^a^, Fy^b^), MNS (M, N, S, s, U, Mi^a^), Diego (Di^a^, Di^b^), Dombrock (Do^a^, Do^b^, Hy, Joa), Colton (Co^a^, Co^b^), Yt (Yt^a^, Yt^b^) and Lutheran (Lu^a^, Lu^b^). HLA genotyping was done by next-generation sequencing (NGS) on a MiSeqDx (Illumina, San Diego, CA, USA), using NGSgo®-Ampx v2 kits, and interpreted with NGSengine® v2.21 (both from GenDX, Utrecht, The Netherlands).

### Statistical analyses

For RBC antigen and HLA allele comparisons in Tables 1, 2 and 3, population-wide proportions were assumed to correspond to the Caucasian prevalence estimates (from the Blood Group Antigen FactsBook^31^ and from the National Marrow Donor Program database^32^), while the HLA G-group allele frequencies of the 90 participants (180 individual HLA alleles) were estimated along with the Clopper-Pearson 95% confidence interval. Z-tests for two proportions were used to test for statistical significance between populational and observed antigen prevalence. A p value inferior to the Bonferroni correction for multiple comparison per antigen group was considered significant.

**Table 1.**
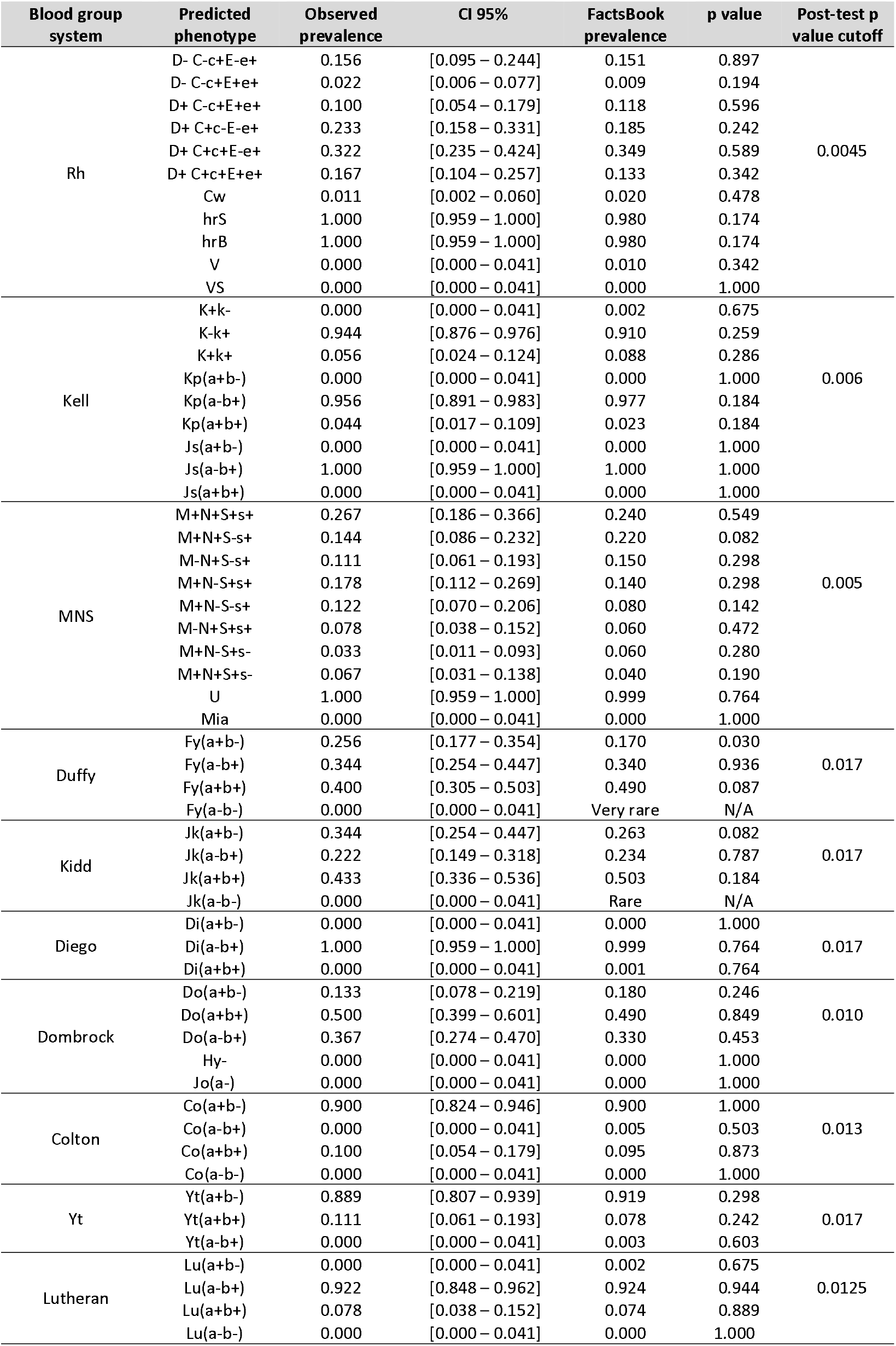
RBC antigen frequencies for the Caucasian convalescent plasma donors compared to reference frequencies

**Table 2.** ABO and RhD blood group distributions for the Caucasian convalescent plasma donors compared to reference frequencies

| Group | Antigen | Observed prevalence | CI 95% | FactsBook prevalence | p value<sup>a</sup> |
| --- | --- | --- | --- | --- | --- |
| All donors<br>(N=90) | A | 0.456 | [0.357 – 0.558] | 0.430 | 0.569 |
|  | AB | 0.089 | [0.046 – 0.166] | 0.040 | **0.018** |
|  | B | 0.100 | [0.054 – 0.179] | 0.090 | 0.741 |
|  | O | 0.356 | [0.264 – 0.458] | 0.440 | 0.110 |
|  | D- | 0.178 | [0.112 – 0.269] | 0.150 | 0.459 |
|  | D+ | 0.822 | [0.731 – 0.888] | 0.850 | 0.459 |
| FY*A/*A<br>individuals<br>(N=23) | A | 0.435 | [0.256 – 0.632] | 0.430 | 0.960 |
|  | AB | 0.130 | [0.045 – 0.321] | 0.040 | **0.028** |
|  | B | 0.130 | [0.045 – 0.321] | 0.090 | 0.503 |
|  | O | 0.304 | [0.156 – 0.509] | 0.440 | 0.190 |

<sup>a</sup> **Bold** p values represents a significant p value of <0.05

**Table 3.** Caucasian convalescent donor HLA allele frequency comparison with the NMDP database for HLA*A, B, C, DRB1 and DQB1. The g notation specifies G-groups corresponding to those presented in Gragert, Loren, Abeer Madbouly, John Freeman, and Martin Maiers. Six-locus high resolution HLA haplotype frequencies derived from mixed-resolution DNA typing for the entire US donor registry. *Human Immunology* 74, n° 10.

| Loci | Alleles | Observed frequency<br>(2N=180) | CI 95% | NMDP frequency | Adjusted p value |
| --- | --- | --- | --- | --- | --- |
| HLA-A | 02:01G | 0,267 | [0.204-0.338] | 0,272 | 1,000 |
|  | 01:01G | 0,122 | [0.078-0.179] | 0,159 | 1,000 |
|  | 03:01G | 0,117 | [0.074-0.173] | 0,140 | 1,000 |
|  | 24:02G | 0,072 | [0.039-0.120] | 0,089 | 1,000 |
|  | 29:02G | 0,061 | [0.031-0.107] | 0,031 | 0,427 |
|  | 11:01G | 0,061 | [0.031-0.107] | 0,059 | 1,000 |
| HLA-B | 44:02G | 0,128 | [0.083-0.186] | 0,087 | 1,000 |
|  | 07:02G | 0,089 | [0.052-0.140] | 0,125 | 1,000 |
|  | 08:01G | 0,083 | [0.047-0.134] | 0,106 | 1,000 |
|  | 44:03 | 0,072 | [0.039-0.120] | 0,044 | 1,000 |
|  | 40:01G | 0,067 | [0.035-0.114] | 0,050 | 1,000 |
|  | 18:01G | 0,061 | [0.031-0.107] | 0,047 | 1,000 |
|  | 35:01G | 0,061 | [0.031-0.107] | 0,058 | 1,000 |
|  | 27:05G | 0,056 | [0.027-0.100] | 0,035 | 1,000 |
| HLA-C | 05:01G | 0,1389 | [0.092-0.198] | 0,083 | 0,109 |
|  | 04:01G | 0,1222 | [0.078-0.179] | 0,115 | 1,000 |
|  | 07:01G | 0,1167 | [0.078-0.179] | 0,155 | 1,000 |
|  | 06:02G | 0,0944 | [0.056-0.147] | 0,095 | 1,000 |
|  | 07:02G | 0,0944 | [0.056-0.147] | 0,137 | 1,000 |
|  | 02:02G | 0,0889 | [0.052-0.140] | 0,047 | 0,138 |
|  | 12:03G | 0,0722 | [0.039-0.120] | 0,056 | 1,000 |
|  | 03:04G | 0,0667 | [0.035-0.114] | 0,074 | 1,000 |
| HLA-DRB1 | 07:01 | 0,178 | [0.125-0.242] | 0,130 | 1,000 |
|  | 01:01 | 0,100 | [0.060-0.153] | 0,085 | 1,000 |
|  | 03:01 | 0,089 | [0.052-0.140] | 0,114 | 1,000 |
|  | 04:01 | 0,089 | [0.052-0.140] | 0,080 | 1,000 |
|  | 13:01g | 0,083 | [0.047-0.134] | 0,000 | 0,000 |
|  | 15:01 | 0,078 | [0.043-0.127] | 0,129 | 0,985 |
|  | 11:01g | 0,056 | [0.027-0.100] | 0,063 | 1,000 |
|  | 14:01g | 0,056 | [0.027-0.100] | 0,027 | 0,497 |
| HLA-DQB1 | 02:01g | 0,211 | [0.154-0.278] | 0,212 | 1,000 |
|  | 03:01g | 0,200 | [0.144-0.266] | 0,196 | 1,000 |
|  | 05:01 | 0,133 | [0.087-0.192] | 0,116 | 1,000 |
|  | 06:03g | 0,089 | [0.052-0.140] | 0,065 | 1,000 |
|  | 06:02 | 0,078 | [0.043-0.127] | 0,128 | 0,621 |
|  | 03:02g | 0,067 | [0.035-0.114] | 0,103 | 1,000 |
|  | 03:03g | 0,056 | [0.027-0.100] | 0,043 | 1,000 |
|  | 05:03g | 0,056 | [0.027-0.100] | 0,029 | 0,407 |

Alleles with less than 10 data points are not shown

For HLA genotype comparison in Tables 4 and 5, individuals from Héma-Québec’s bone marrow donor registry were used as reference. In total, 1370 registered individuals typed in high-resolution, from the Montréal and Montérégie regions and of self-reported Caucasian ethnicity, were selected to match with the characteristics of the studied convalescent cohort. Allele frequencies were calculated using the GENE[RATE] tool for HLA-A, B, C, DRB1, DQB1, and DPB1^33^. HLA allelic frequencies were used with a modification of the hierfstat package to calculate the genetic distance (Latter) globally and for each locus between the subjects and the reference population^34,35^. Standardized residuals of the cohort subjects were calculated to identify alleles with significant differences between the subjects and the registry. Residuals were calculated by considering the subjects’ allele frequencies as the independent variable and the registry frequency as the dependant variable for each locus independently. Frequencies were deemed different at or above a difference of 3 absolute standardized residual. Statistical analyses were performed using the free statistical software R^36^.

**Table 4.** Genetic distance comparison between Caucasian convalescent donor HLA allele frequencies (2N=180) and Héma-Québec bone marrow donor registry (2N=2740) for HLA*A, B, C, DPB1, DQB1 and DRB1, per loci, and globally.

| Locus | A | B | C | DPB1 | DQB1 | DRB1 | Global |
| --- | --- | --- | --- | --- | --- | --- | --- |
| Genetic Distance | 0,0017 | 0,0050 | 0,0045 | 0,0018 | 0,0021 | 0,0040 | 0.0033 |

**Table 5.** HLA alleles identified as significantly over- or underrepresented from pairwise comparisons between the convalescent cohort (2N=180) and Héma-Québec bone marrow donor registry (2N=2740), and their potential clinical significance.

| Alleles | Frequency |  | Standardised residuals | Significance and references |
| --- | --- | --- | --- | --- |
|  | Cohort | Registry |  |  |
| A*01:01 | 0,1222 | 0,1456 | -3,3570539* | Associated with high risk<sup>45</sup>, mortality<sup>46</sup> |
| B*44:02 | 0,1278 | 0,0772 | 5,15830039* | B*44 More susceptible to infection<sup>26</sup> |
| B*51:01 | 0,0222 | 0,0640 | -3,6358415* | B*51 non-significantly correlates with infection susceptibility<sup>26</sup> |
| C*05:01 | 0,1389 | 0,0848 | 4,11569996* | Risk of death<sup>27</sup> |
| DPB1*04:01 | 0,4167 | 0,4048 | 3,39285576* | Direct association with severity<sup>48</sup> |
| DPB1*04:02 | 0,0833 | 0,1128 | -3,7313073* | No data reported |
| DRB1*04:01 | 0,0889 | 0,0488 | 3,26507272* | Increased in severe vs. asymptomatic<sup>49</sup> |
| DRB1*07:01 | 0,1778 | 0,1397 | 3,85103473* | No data reported |

\*Standardised residuals $\geq 3$ or $\leq -3$ are considered significant

## Results

### Red blood cell genotype frequencies

Genotype frequencies for Rh, Kell, MNS, Duffy, Kidd, Diego, Dombrock, Colton, Yt and Lutheran blood groups were determined in each individual, and the resulting predicted phenotypes were compared to the expected Caucasian reference frequencies. The FY*A/*A genotype (Fy(a+b-) predicted phenotype) appears to be overrepresented (non-significant, p=0.030) in our convalescent cohort compared to expected frequencies (0.256 vs 0.170, respectively), as presented in Table 1. Incidentally, FY*A/*B (Fy(a+b+) predicted phenotype) individuals appear to be trending towards a decreased frequency of 0.400 compared to the expected 0.490 (Table 1), although the trend is not significant (p=0.087). Overall, no antigen group combinations deviated significantly from the expected frequencies.

### ABO and RhD phenotyping

ABO and RhD phenotypes were determined for each convalescent individual and compared to the expected Caucasian reference frequencies for the entire cohort (N=90) and within FY*A/A individuals (N=23), given its non-significant but trending towards overrepresentation (Table 1). Table 2 presenting the ABO phenotyping analysis for the cohort allowed for the identification of a significant (p=0.0178) 2.2x increase for the AB group compared to reference frequencies (0.089 vs 0.040, respectively). While non-significant (p=0.110), the O group trends towards a 0.8x underrepresentation within convalescent individuals compared to expected frequencies (0.356 vs 0.440, respectively). Interestingly, a significant (p=0.028) 3.3x AB group overrepresentation was also found within FY*A individuals (Table 2, FY*A/*A). An apparent but non-significant decrease in O group individuals can be observed within the FY*A/*A individuals versus the reference frequency (0.304 vs 0.440). No other trend or significant observation was made regarding A and B groups, nor regarding RhD expression.

### HLA allele frequency comparisons

HLA typing was done by NGS in the Caucasian convalescent donors’ cohort and individual allele frequencies were determined. The convalescent cohort allele frequencies (2N=180) were first compared to the NMDP registry frequencies for HLA-A, B, C, DRB1 and DQB1 for the most frequent alleles (Table 3), and no significant differences were identified in the most common alleles in the cohort. The convalescent cohort frequencies for HLA-A, B, C, DRB1, DQB1 and DPB1 were then compared to the Héma-Québec Stem Cell Donor Registry frequencies for Caucasians in the same geographical region (2N=2740). The genetic distance between the convalescent donors’ cohort and the Héma-Québec Registry was calculated for all loci together and on a per-locus basis for HLA-A, B, C, DRB1, DQB1, and DPB1 (Table 4). There was no comparative measurement available for the genetic distance, however the distance is low in all loci compared. Standardized residual analysis was conducted (Table 5 and Figure 1) and led to the identification of alleles that were significantly different between the convalescent cohort (N=90 individuals) and the Héma-Québec Registry (N=1370 individuals). HLA-B*44:02, C*05:01, DPB1*04:01, DRB1*04:01 and DRB1*07:01 were significantly overrepresented within our cohort, while A*01:01, B51:01 and DPB1*04:02 were significantly underrepresented. Finally, the alleles identified as significantly different were used to search previously published data for suspected associations with the SARS-CoV-2 virus infection and COVID-19 disease characteristics; these results are presented in Table 5. Of note, for two of the eight alleles that we have identified (DPB1*04:02 and DRB1*07:01), there was no data in the literature for comparison.

**Figure 1.**
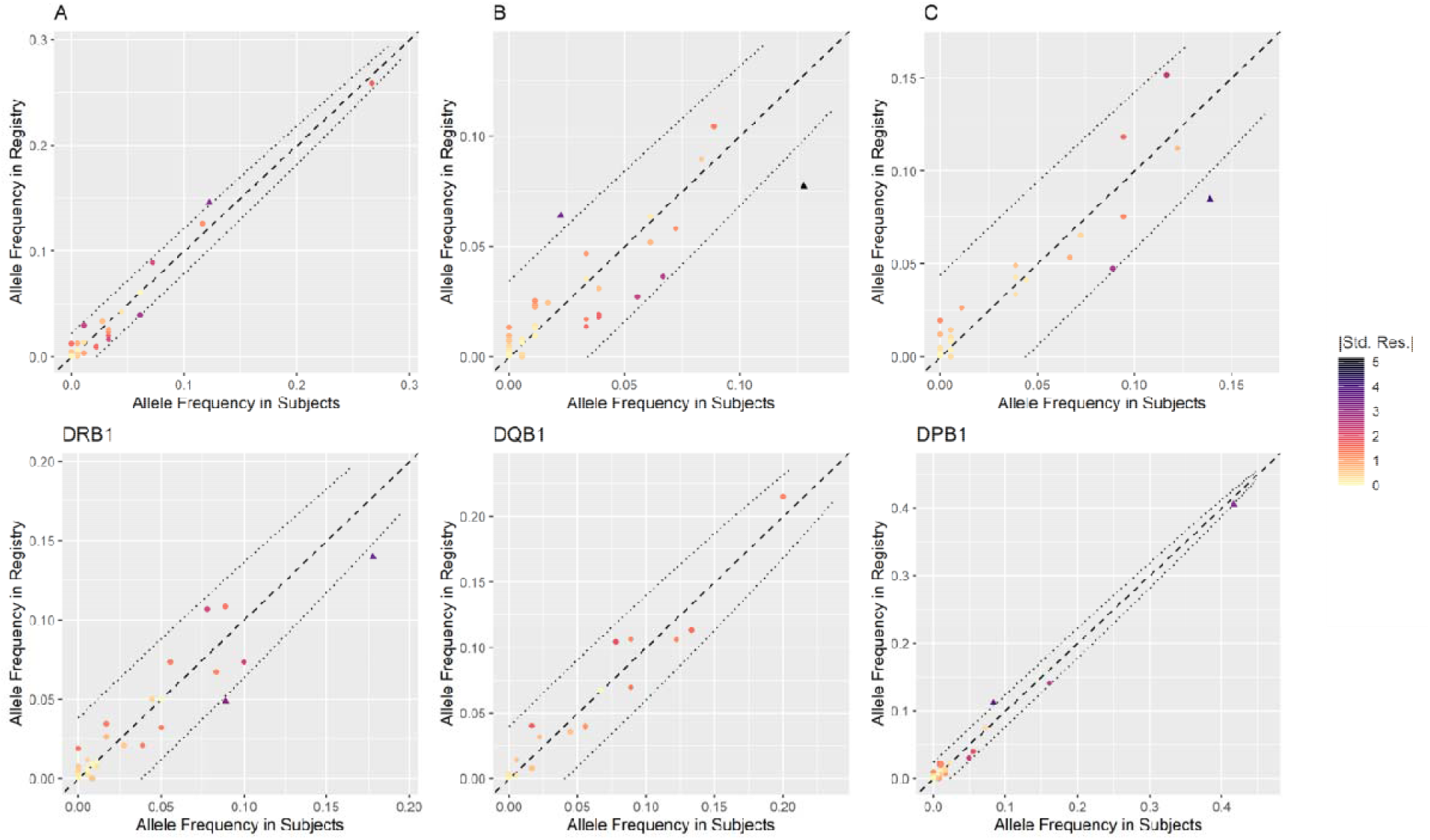
HLA Allelic frequencies in the convalescent cohort and the Héma-Québec bone marrow donor registry. For each locus, the cohort allele frequencies (horizontal axis) are plotted against the registry frequencies (vertical axis). The dashed line represents identical expression in the cohort and registry. The color of the alleles represents the absolute standardized residuals (|Std. Res.|) to the identity line. The dotted lines are the thresholds of 3 |Std. Res.| away from the identity line.

## Discussion

Our study took a deeper look into the RBC and HLA characteristics of a COVID-19 recovered and non-hospitalized cohort enrolled in the CONCOR-1 convalescent plasma study. We used ABO and RhD automated blood donor testing, and RBC and HLA high throughput genotyping platforms to determine the existence of potential trends regarding the frequencies of ABO, RhD, 37 other RBC antigens and HLA genotypes within the cohort, compared to reference Caucasian populations from textbooks, public databases, and our local bone marrow donor registry. A significant AB blood group overrepresentation was identified, as well as a non-significant trend in FY*A/A individuals. These results suggest a possible involvement of ABO and Duffy red blood cell antigens in SARS-CoV-2 susceptibility and COVID-19 severity, as all these individuals contracted the virus, yet only had mild symptoms, and cleared the infection without hospitalization.

The case of HLA association with disease susceptibility and severity is more complex. Overall, the genetic distance calculated from HLA allele frequencies is low (below 0.01) and suggests the cohort is similar to the reference population chosen. When looking at allele-level frequencies, eight HLA alleles were identified as potential markers between the convalescent cohort and a stem cell registry from the same geographical region by the standardized residuals analysis. We have also considered using 2×2 contingency tables along with Chi Squared tests to evaluate individual allelic differences. While some differences were significant after multiple testing corrections (HLA-A*68:02, B*39:06, B*51:01, B*52:01, C*12:02, C*14:02, DPB1*15:01, and DRB1*13:03), they involved less frequent alleles in the Héma-Québec Registry that were mostly absent from the cohort. Of these alleles, only HLA-B*51:01 was observed in the convalescent cohort, but at a lower frequency than in the Registry. Interestingly, this allele was also significant in the standardized residual analysis. One of the limitations of Chi Squared tests is the need for sufficient observed values (>5), which is not respected in these cases and prevents us from drawing conclusions from this analysis. Further studies and more data will be required to draw conclusion from these hypothetical associations.

One of the limitations of this study concerns the lack of a more diverse stratification of disease severity and the limited sample size for COVID-19 affected individuals. Indeed, our study lacks blood group and HLA data from hospitalized, deceased and asymptomatic COVID-19 patients, especially given that there could be a significant link between ABO and severity^37^. Additionally, while the historical ABO frequencies of Quebec Caucasian blood donors (internal data) matched that of FactsBook, such unbiased information about the frequencies of other RBC antigens is not currently available, hence the use of FactsBook Caucasian reference frequencies. Our study also does not directly address the major RBC and HLA antigen frequency differences between ethnicities. While our convalescent plasma donation program reflected our donor pool^38^, the low number of non-Caucasian individuals was insufficient to conduct statistical analysis, which is unfortunate given the importance of understanding the disproportionate impact of COVID-19 on minorities^5,29^. Nonetheless, the identification of a potential overrepresentation of FY*A/*A within Caucasian COVID-19 convalescent individuals and the potential implication of the Duffy blood group could have an impact on future research directions. The trend towards overrepresentation of the Fy(a+b-) predicted phenotype among our COVID-19 convalescent cohort could be explained by the absence of the Fy^b^ antigen, since no significant difference was observed when comparing Fy(a+b+) and Fy(a-b+). In individuals of African descent, the Fy(a-b-) phenotype is caused by a GATA box mutation upstream of the *FY* gene silencing Fy^b^ expression in RBCs^39^. Given that 67% of African Americans (AA) are Duffy null^40^, and that Duffy null patients have an increased mortality rate as a consequence of acute lung injury^4^, some groups have already hypothesized a role for Duffy in COVID-19 AA individuals^40^. We therefore suggest that Duffy allele identification might be used to select individuals at-risk for COVID-19 complications for further research on associations between COVID-19 and RBC antigens. The involvement of ABO blood groups in COVID-19 has previously been described^13,41^. The mechanism underlying the association remains elusive, but could be related to circulating natural anti-A and anti-B antibodies, or a low-efficiency furin cleavage in O-group individuals^9^. A significant overrepresentation of the AB group, and the non-significant but trend towards underrepresentation of the O group within our cohort, appear to be in agreement with other groups’ suggestion that O individuals could be less susceptible to SARS-CoV-2 infection^11,12^, given that infection was confirmed in every individual in our study. Our sample size does not allow us to draw conclusions as to whether individuals from the AB group are more susceptible to infection and more efficient at clearing infection without hospitalization, or whether this bias is a consequence of the trend towards underrepresentation of O individuals who are less susceptible to infection. It would be interesting to extend our observations within larger cohort analyses that include severely affected patients.

The involvement of HLA alleles in the disease outcome of COVID-19 patients is progressively getting clearer^42,43^. Various HLA alleles have shown high binding affinity to SARS-CoV-2 peptides,^18,44^ and trends have been observed in many populations^22,23^. While our sample size is limited, eight alleles were identified as significantly different between the studied cohort and matched individuals from the Héma-Québec Stem Cell Donor Registry, for which we already had high resolution HLA information. Two of these eight HLA alleles have not been previously identified for their association with COVID-19 in the literature: DPB1*04:02 and DRB1*07:01. The underrepresentation of HLA-A*01:01 is in agreement with its suggested association with high risk and mortality in COVID-19 individuals^45,46^, and the overrepresentation of HLA-B*44:02 is compatible with an increased susceptibility to infection^26^. Interestingly, A*01:01, B*44:02 and B*51:01 were predicted as weak binder of SARS-CoV-2 peptides, and none of the other alleles we identified were found to be strong peptide binders^47^. Given that we found A*01:01 and B51:01 to be underrepresented and B*44:02 overrepresented in our cohort, it is difficult to establish a relationship between these data without considering complete haplotypes or other disease severity groups. The other associations are inconclusive, but should be considered in larger cohorts. Overall, more data is required from as many diverse populations as possible in order to develop a comprehensive view, and to have data readily available for emerging variants and infection waves.

Altogether, we provide additional information regarding the role of RBC antigens and HLA in SARS-CoV-2 susceptibility, and consequential COVID-19 susceptibility, severity, resolution and long-term clinical consequences. More research needs to be done to get a better understanding of potentially at-risk populations, and for the identification of molecular pathways of this virus and its variants.

## Data Availability

None.

## Acknowledgements

The authors are grateful to the convalescent plasma donors who participated in this study and the Héma-Québec team involved in convalescent donor recruitment, sample collection, as well as the technologists who performed the phenotyping and genotyping workup. The authors thank Jean-Francois Leblanc and Marie-Eve Rhéaume for revising the manuscript. The authors are also thankful for healthcare workers’ dedication worldwide.

P.T. and W.L. designed the research and wrote the manuscript. P.T., L.R. and M-C.C. supervised the research. J.P., L.R., W.L. and J.C.Y. acquired the data. P.T., J.P., L.R., W.L., N.B., A.L. and J.C.Y. analysed the data. P.T., L.R., W.L., G.A.L, N.B., M-C.C and J.C.Y. reviewed and edited the manuscript.

